# Control of Center of Mass Motion during Walking Predicts Gait and Balance in People with Incomplete Spinal Cord Injury

**DOI:** 10.1101/2023.01.19.23284492

**Authors:** Shamali Dusane, Anna Shafer, Wendy L. Ochs, Tara Cornwell, Heather Henderson, Kwang-Youn A. Kim, Keith E. Gordon

## Abstract

**Background:** There is evidence that ambulatory people with incomplete spinal cord injury (iSCI) have an impaired ability to control lateral motion of their whole-body center of mass (COM) during walking. This impairment is believed to contribute to functional deficits in gait and balance, however that relationship is unclear. Thus, this cross-sectional study examines the relationship between the ability to control lateral COM motion during walking and functional measures of gait and balance in people with iSCI.

**Methods:** We assessed the ability to control lateral COM motion during walking and conducted clinical gait and balance outcome measures on twenty ambulatory adults with chronic iSCI (C1-T10 injury, American Spinal Injury Association Impairment Scale C or D). To assess their ability to control lateral COM motion, participants performed three treadmill walking trials. During each trial, real-time lateral COM position and a target lane were projected on the treadmill. Participants were instructed to keep their lateral COM position within the lane. If successful, an automated control algorithm progressively reduced the lane width, making the task more challenging. If unsuccessful, the lane width increased. The adaptive lane width was designed to challenge each participant’s maximum capacity to control lateral COM motion during walking. To quantify control of lateral COM motion, we calculated lateral COM excursion during each gait cycle and then identified the minimum lateral COM excursion occurring during five consecutive gait cycles. Our clinical outcome measures were Berg Balance Scale (BBS), Timed Up and Go test (TUG), 10-Meter Walk Test (10MWT) and Functional Gait Assessment (FGA). We used a Spearman correlation analysis (ρ) to examine the relationship between minimum lateral COM excursion and clinical measures.

**Results:** Minimum lateral COM excursion had significant moderate correlations with BBS (ρ=−0.54, p=0.014), TUG (ρ=0.59, p=0.007), 10MWT-preferred (ρ=−0.59, p=0.006), and FGA (ρ=−0.59, p=0.007) and a significant strong correlation with 10MWT-fast (ρ=−0.68, p=0.001).

**Conclusion:** Control of lateral COM motion during walking predicts a wide range of clinical gait and balance measures in people with iSCI. This finding suggests the ability to control lateral COM motion during walking could be a contributing factor to gait and balance in people with iSCI.

## Introduction

The requirements of the nervous system to actively control mediolateral motion of the whole-body center of mass (COM) during walking are substantial in comparison to other planes of motion that benefit from stabilizing body mechanics [1–3]. Growing evidence suggests that ambulatory people with incomplete spinal cord injury (iSCI) have considerable challenges controlling their lateral COM motions during walking. This includes difficulty arresting lateral motion after a walking maneuver [4, 5], impaired mediolateral foot placement [6], limited ability to increase lateral margins of stability following a perturbation [7], and sizeable metabolic energy cost to stabilize lateral motion during walking [8]. Many of the cautious gait patterns observed in people with iSCI (e.g., slower walking speeds, wider steps, shorter steps, more time in double support [6, 9–12]) have been suggested to be compensatory mechanisms that proactively aid in COM control during walking [9, 10, 12].

Studies in populations without neurologic injuries have found that the control of mediolateral COM motion is critical for maintaining dynamic balance [13] and creating walking stability during directional changes [14]. Almost all activities of daily living involve directional changes or turning maneuvers that require reorientation of the body in the anticipated direction of travel [15]. Therefore, the ability to control lateral COM motion during walking may be a skill fundamental to functional gait and balance. If this relationship is supported, interventions that directly aim to improve the ability to control lateral COM motion may translate to improvements in gait and balance. The first step is to identify if the ability to control lateral COM motion during walking is related to functional gait and balance in people with iSCI.

The ability to control lateral COM motion during walking could be related to several functional measures of gait and balance. We selected four clinical outcome measures, the Functional Gait Assessment (FGA), the Timed Up and Go test (TUG), the 10-meter walk test (10MWT), and the Berg Balance Scale (BBS), that collectively would provide insights into the relationship between the ability to control lateral COM motion during walking and walking balance, walking speed, and postural balance, respectively. People with iSCI have impaired abilities to control lateral motion during walking [4–7], which may reduce functional walking balance on tasks requiring turns and change of gait speed. In the current study, we used the TUG [16–18] and the FGA [19–21] to examine walking balance. Both these tests include turns and changes in walking speed. People with iSCI often select cautious gait patterns, including walking at slower speeds [9, 10, 12], that reduce COM velocity. Slower COM motions are believed to enhance gait stability by decreasing perturbation intensities [9, 12]. Thus, it seems like that there could also be a relationship between the ability to control COM motion during walking and walking speed. We conducted the 10MWT, a widely used and recommended measure to assess gait speed [22–25], and analyzed relationship of preferred and fast 10MWT speeds with the ability to control lateral COM motion. Finally, the BBS evaluates postural balance but has strong correlations with walking ability in people with iSCI [26–29], suggesting that similar mechanisms may be responsible for controlling COM dynamics during walking and standing. In the current study, we used the BBS to examine if the ability to control lateral COM motion during walking is related to postural balance. While these measures provide an overview of functional gait and balance, it should be noted that they do not provide a direct method to assess the ability to control lateral COM motion during walking.

To quantify the ability to control lateral COM motion during walking, we developed a laboratory-based assessment. During this assessment, a target walking lane and the real-time mediolateral position of the participant’s COM are projected on the walking surface of an oversized treadmill. Participants are instructed to maintain their mediolateral COM position within the projected lane during walking. If participants maintain their COM within the target lane, the lane width is progressively decreased, increasing the challenge of the walking task. This external visual feedback encouraged participants to try their best to control their COM motion during forward walking. To quantify the participants’ capacity to control their lateral COM motion, we performed a post-hoc kinematic analysis to identify the minimum lateral COM excursion occurring during five consecutive gait cycles.

The purpose of this study was to evaluate the relationship between the ability to control lateral COM motion during walking and validated clinical gait and balance measures commonly used to assess ambulatory people with iSCI. We hypothesized that the ability to control lateral COM motion during walking would be predictive of clinical gait and balance outcome measure scores.

## Materials and methods

### Participants

Twenty adults with chronic incomplete spinal cord injury participated in this cross-sectional study. All participants had spinal cord injuries between C1-T10 and were classified as C or D on the American Spinal Injury Association Impairment Scale (AIS). Our inclusion criteria were the following: age between 18 to 80 years, more than 6 months post-incomplete spinal cord injury, medically stable, and able to ambulate 10m without physical assistance or assistive devices such as an ankle foot orthosis (AFO), cane or rolling walker. Our exclusion criteria were the following: excessive spasticity in the lower limbs (> 3 on the Modified Ashworth Scale), unable to tolerate 10 minutes of standing, presence of severe cardiovascular and pulmonary disease, unhealed decubiti or other skin compromise, history of recurrent fractures or known orthopedic problems in the lower extremities, concomitant central or peripheral neurological injury, unable to provide informed consent due to cognitive impairments, enrolled in concurrent physical therapy or research involving locomotor training, and use of braces/orthotics crossing the knee joint. This study was conducted at the Human Agility Laboratory, Physical Therapy and Human Movement Sciences, Northwestern University Feinberg School of Medicine. The study protocol was approved by the Institutional Review Boards at Northwestern University and the Edward Hines Jr. Veterans Affairs Hospital. All participants provided informed written consent prior to enrollment in the study.

### Experimental Setup

To assess participants’ ability to control their lateral COM motion during walking, we recorded kinematic data as participants walked on an oversized treadmill, walking surface 2.6 × 1.4 m, (TuffTread, Willis, TX) while receiving visual feedback about their lateral COM position. For safety, participants wore a trunk harness attached to passive overhead support that did not provide bodyweight support (Aretech, Ashburn, VA). The harness straps were adjusted to allow participants unrestricted lateral travel across the treadmill. During treadmill walking, participants were not allowed to use any assistive devices (canes, walkers, handrails) except for any passive ankle-foot orthoses they would typically wear during community ambulation. In the event of a loss of balance, a physical therapist providing standby assistance would give manual support as necessary to allow the participant to regain balance and continue walking. During the assessment of participants’ ability to control their lateral COM motion, we only analyzed walking periods when no manual assistance was provided.

During treadmill walking, we used a 12-camera motion capture system (Qualisys, Gothenburg Sweden) to collect 3D coordinates of 19 reflective markers placed on the pelvis and lower limbs at 100 Hz. Markers were placed at the following locations: S2 vertebrae and bilaterally on each sacroiliac joint, greater trochanter, anterior superior iliac spine, highest point of the iliac crest, lateral malleolus, calcaneus, and the 2^nd^, 3^rd^, and 5^th^ metatarsals.

Participants were given visual feedback of their lateral COM position to challenge them to minimize their lateral motion during treadmill walking. Specifically, their real-time mediolateral COM position was represented by a white line projected along the length of the treadmill surface using a short throw projector mounted on the wall alongside the treadmill (Hitachi, Tokyo, Japan). The lateral COM position was calculated using real-time 3D locations of the pelvis markers that were streamed to a custom-programmed control algorithm (LabVIEW, National Instruments, Austin, TX). The control algorithm calculated mediolateral COM position as the midpoint between the two greater trochanter markers [30] and transformed the data into the treadmill coordinate system for display.

Additionally, lateral boundary targets for COM position (“target lane”) were projected on the treadmill (Figure 1). To maximally challenge participants to minimize their lateral COM motion during walking, the control algorithm systematically adjusted the width of the target lane based on how successful the participant was at maintaining their lateral COM position within the green target lane (Figure 1). During walking, if the COM moved outside the target lane, that area outside the target turned red to provide an immediate visual cue to return to the green target lane. At the beginning of the first walking trial, the initial lane width was set to 200mm. Once the assessment began, the control algorithm made a 10mm step change in the lane width based on the following logic: if the participant maintained their lateral COM position in the lane for 1.5 consecutive meters of forward walking, the lane width was decreased by 10mm. If the participant walked for 3 meters without maintaining COM position within the lane for at least 1.5 consecutive meters, the lane width increased by 10mm. A minimum lane width was set at 5mm. Consecutive walking assessments started at the lane width achieved at the end of the previous trial (e.g., the starting lane width for the second assessment was equal to the ending lane width of the first assessment). The algorithm thresholds were established in pilot testing prior to the current study to minimize walking time yet converge on the smallest lane width participants could maintain.

**Figure 1.**
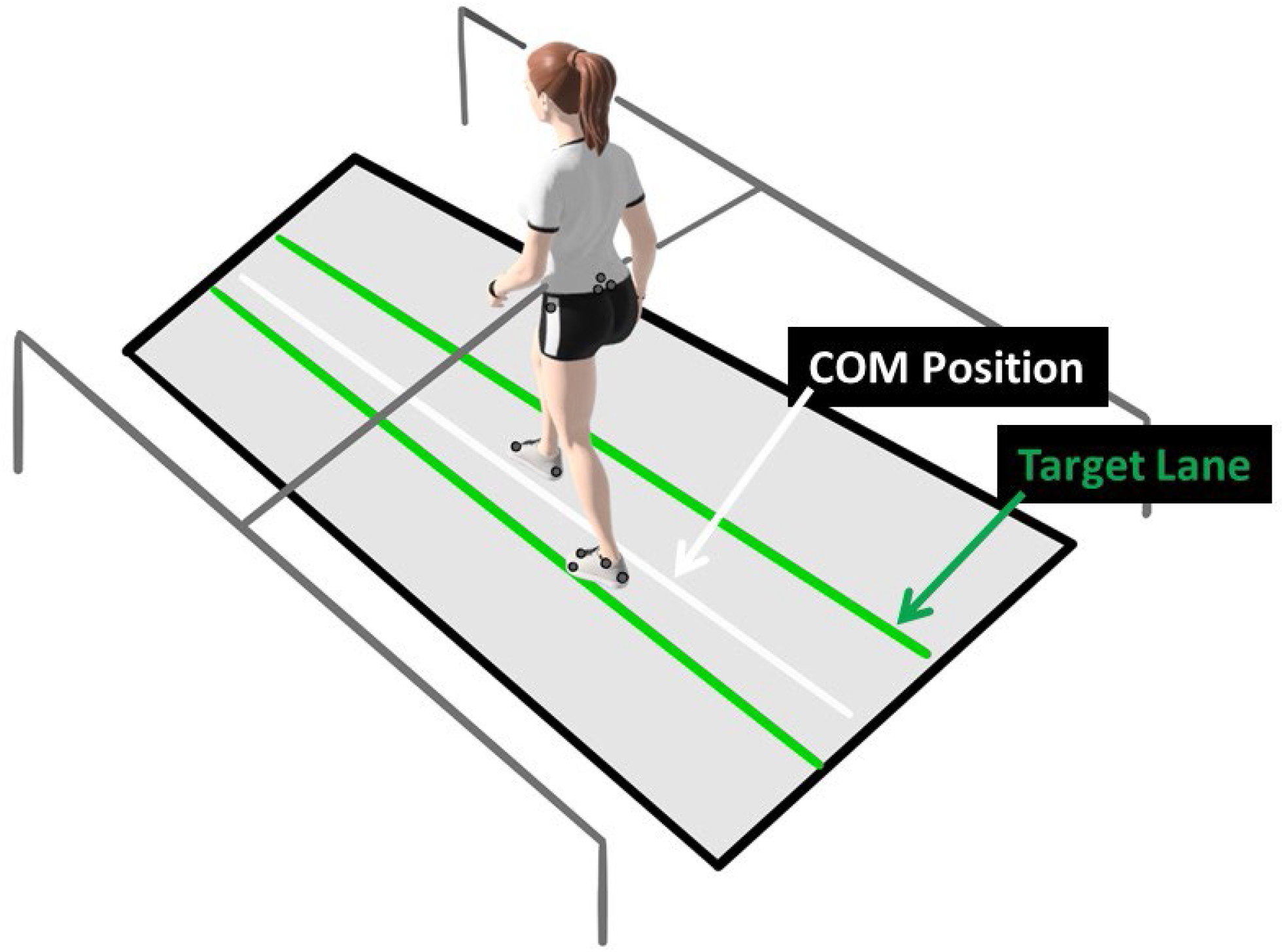
Experimental set-up. The laboratory balance assessment is performed three times on an oversized treadmill to test an individual’s lateral center of mass (COM) control. The real-time mediolateral COM position of the participant is projected on the treadmill through a white line. Participants are instructed to walk at their preferred treadmill speed and to do their best to keep the white line within the green target lane. If successful, the target lane width is progressively decreased. If not, the area outside of the target lane (to either the left or right) changes to red, providing an immediate visual cue that the participant has made an error and should try to return the white line to the projected increased lane.

### Clinical outcome measures

#### Timed Up and Go test (TUG)

The TUG is a valid and reliable measure for functional mobility, balance, and fall-risk in people with iSCI [16, 17]. It is a widely used and recommended screening tool for prediction of fall-risk [31–33]. The participant is asked to rise from the chair, walk 3 meters safely, turn around, walk back, and sit down on the chair. The TUG score is the time recorded from when participants rise from the chair until they sit down again.

#### Functional Gait Assessment (FGA)

The FGA is a valid and reliable test that measures balance and gait functions in people with iSCI [20, 21]. It is a modification of the Dynamic Gait Index [19] consisting of 10 items scored on a four-point ordinal scale ranging from 0 to 3, such that 0 indicates severe impairment and 3 indicates normal/no impairment with a total score of 30. The clinical practice guideline recommends usage of FGA as a core outcome for patients with neurological conditions [34].

#### Berg Balance Scale (BBS)

The BBS is a 14-item scale, widely used, valid, and reliable measure for balance assessment of people with iSCI during predetermined tasks performed in daily living [27–29]. Each item is scored on a five-point ordinal scale ranging from 0 to 4, such that 0 signifies the lowest level of function and 4 signifies the highest level of function. The maximum total score is 56, with higher total scores indicating better balance [35]. The BBS has also been useful as a screening tool to predict risk of falls beyond cut-off scores [36–38].

#### 10-Meter Walk Test (10MWT)

The 10MWT has been found to be a valid and reliable test to measure overground walking speed, both preferred and fast, in people with iSCI [17, 39–41]. It is a recommended measure for assessment of gait speed among neurological populations [24, 25].

### Protocol

All participants first underwent a clinical assessment. A licensed physical therapist collected demographic information (age, gender, date of birth), date of spinal cord injury, level of spinal cord injury, cause of spinal cord injury, current and past medical history, current medications, current ambulatory ability in the home and community (including the use of any assistive devices), self-reported number of falls in the past year. The physical therapist then collected four clinical outcome measures: BBS, TUG, FGA, and 10MWT at preferred and fast speeds.

Next, participants performed the treadmill walking portion of the experiment that was used to assess their ability to control their lateral COM motion. The participant’s preferred treadmill walking speed was identified through a staircase method of increasing and decreasing the treadmill speed until the participant’s desired speed is confirmed through verbal feedback. Participants were given several minutes to accommodate to walking on the treadmill at this preferred speed.

With the treadmill stopped, participants were then given detailed instructions about the assessment to be performed. The projector used to display the participant’s real-time lateral COM position and a target lane was turned on. Participants were instructed to make some small movements to their left and right so that they understood that the side-to-side movement of their body controlled the position of the white line being projected on the treadmill. Participants were asked to perform three walking trials of 21 m each. Participants were instructed to do their best to maintain the white line representing the midline of their body within the target lane. They were also told that if they were successful, the width of the target lane would be progressively reduced. Once the participant understood the instructions, the treadmill was started, and participants were given time to reach steady state before the assessment began. At the end of the 21 m assessment, the treadmill was stopped, and participants were given time to rest as needed. Then two more 21 m assessments, separated by a rest break, were performed.

### Data Analysis and Processing

Data from all treadmill walking assessments were examined and used to estimate participants’ ability to control their lateral COM motion during treadmill walking. Kinematic marker data was processed using Visual3D (C-Motion, Germantown, MD) and a custom MATLAB (Mathworks, Natick, MA) program. Marker data was gap-filled and low-pass filtered (Butterworth, 6 Hz cut-off frequency). Time of initial foot contact (IC) and toe-off (TO) events were identified for each step based on maximum and minimum fore-aft positions of the calcaneus and 2nd metatarsal markers, respectively. Mediolateral COM position was calculated in Visual3D as the center of the Visual3D model’s pelvis. We then calculated the lateral COM excursion for each gait cycle. Finally, from all gait cycles, we identified the five consecutive gait cycles that produced the smallest average lateral COM excursion. This value, the minimum lateral COM excursion over five consecutive gait cycles, was used to represent each participant’s ability to control their lateral COM motion during forward walking.

### Statistical analysis

Descriptive variables, scores of clinical outcome measures, and minimum lateral COM excursion were reported as mean (standard deviation (SD)). The Shapiro-Wilk test for normality was used to confirm the assumption of normality of clinical balance measures and minimum lateral COM excursion. To evaluate the relationship between the ability to control lateral center of mass motion during walking and clinical outcome measures, a Spearman correlation analysis was performed between minimum lateral COM excursion and the following clinical outcome measures: TUG, FGA, BBS and preferred and fast 10MWT. Spearman’s correlation coefficients (ρ) used for all the measures was interpreted as follows: >0.70 as strong, 0.50-0.70 as moderate, 0.30-0.50 as weak [42]. All statistical analyses were performed using SPSS (Version 24, SPSS Inc., Chicago, IL, USA) with α=0.05.

## Results

Twenty people with iSCI participated in our study (mean age of 52.9±18.06 years). Participant characteristics are reported in Table 1. All participants were able to successfully complete the walking assessments.

**Table 1.** Participant characteristics

| **Demographics (n=20)** | **Results** |
| --- | --- |
| Age, mean (SD), years | 52.9 (18.06) |
| Sex (number (%)) |  |
| Male | 14 (70%) |
| Female | 6 (30%) |
| Height, mean (SD), inches | 67.95 (2.94) |
| Years post SCI | 7.95 (9.06) |
| Level of injury (number) |  |
| Cervical | 13 |
| Thoracic | 7 |
| Mechanism of injury (number) |  |
| Traumatic | 15/20 |
| Non-traumatic | 5/20 |
| Self-selected Walking Index for Spinal Cord Injury (WISCI II) | 17.65 (2.74) |
| Lower Extremity Motor Score (LEMS) | 43.85 (4.69) |
| Walking device |  |
| Rolling walker | 1/20 |
| Cane | 10/20 |
| None | 9/20 |
| Brace usage |  |
| Using brace | 4/20 |
| Not using brace | 16/20 |

Minimum lateral COM excursion significantly correlated with all clinical outcome measures we examined. There was a moderate, positive correlation between minimum lateral COM excursion and TUG time (ρ=0.59, p=0.007) (Figure 2a). There was a moderate, negative correlation between minimum lateral COM excursion and FGA score (ρ= −0.59, p=0.007) (Figure 2b) and BBS score (ρ= −0.54, p=0.014) (Figure 2c). The minimum lateral COM excursion had a strong, negative correlation with fast 10MWT speed (ρ= −0.68; p=0.001) (Figure 3a) and a moderate, negative correlation with preferred 10MWT speed (ρ= −0.59; p=0.006) (Figure 3b).

**Figure 2.**
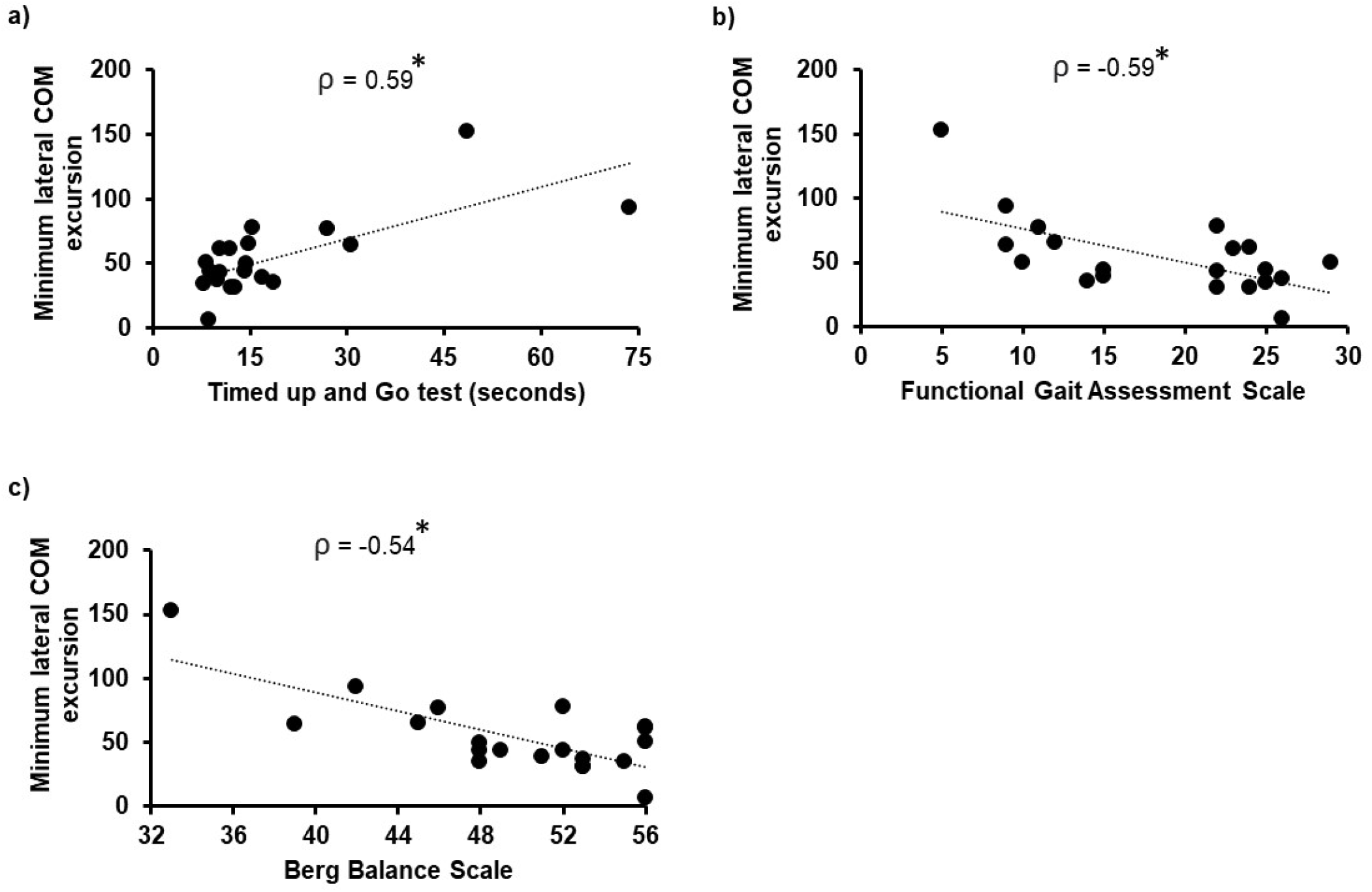
Scatterplots of minimum lateral COM excursion with a) Timed Up and Go test, b) Functional Gait Assessment Scale, and c) Berg Balance Scale. Each circle represents one participant. *indicates p <0.05.

**Figure 3.**
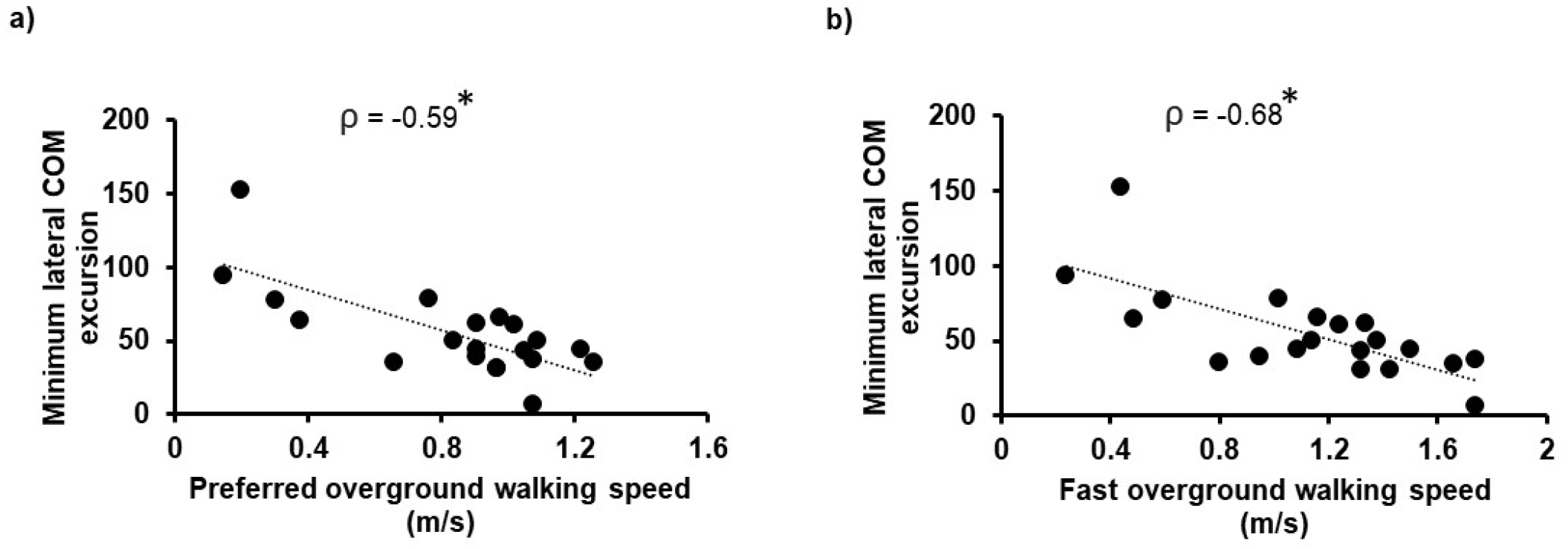
Scatterplots of minimum lateral COM excursion with a) preferred overground walking speed as measured by the 10MWT and b) fast overground walking speed as measured by the 10MWT. Each circle represents one participant. *indicates p <0.05.

## Discussion

In support of our hypothesis, we found that for ambulatory adults with iSCI, the ability to control their lateral COM motion during walking was moderately to strongly correlated with clinical outcome measures related to walking balance, walking speed, and postural balance.

The ability to control lateral COM motion during walking was found to be moderately correlated with our two clinical measures of walking balance, the TUG and FGA. Smaller minimum lateral COM excursions were associated with shorter TUG times and higher scores on the FGA, both indicating greater walking ability. A moderate correlation indicated that both (clinical measure and lateral COM excursion) measure a similar construct i.e., walking balance. Both the TUG and FGA involve walking and maneuvering with a pivot turn that challenges functional walking balance. Specifically, the FGA provides a range of gait activities (change in gait speed, head turns, narrow base of support walking) that require complex stabilizing strategies and postural adjustments. Whereas the TUG requires motor planning and capacity to anticipate transitioning from one motor task to another in a particular sequence [43]. Due to their sensorimotor deficits, reduced muscle strength, impaired proprioception and deficits in balance and coordination demonstrate, people with iSCI have difficulty with safely performing complex maneuvers during walking. These impairments challenge dynamic balance and their ability to make anticipatory changes during walking. Our finding that individuals with the poorest ability to control their lateral COM excursion also had the lowest scores of FGA and took the longest time to perform TUG is built on previous findings indicating that people with iSCI have impaired control of lateral motion during walking [4–7].

Similar to TUG and FGA, minimum lateral COM excursion showed a moderate, negative correlation with BBS score. Specifically individuals with greater BBS scores, which indicate greater postural control and balance, demonstrated a better ability to control their lateral COM excursion during walking. The BBS is often referred to as a “gold standard” because it is one of the most widely used, valid, and reliable clinical measures for assessing balance and postural control [28, 29, 36, 44, 45]. Since scoring for BBS is based on how well the participant performs balance challenging task that are performed in daily life, it is a good indicator of functional balance (including static and dynamic) and used for assessment of fall-risk in several populations [46–49]. Previous studies examining the relationship between balance measured by BBS and walking ability in neurological population found that BBS score is strong predictor of walking ability (home and community ambulation) among people with stroke [50–52]. BBS score is also found to correlate with walking performance in people with SCI [26] demonstrating a strong relationship between walking function and balance. Our results support this relationship such that people with iSCI who had the best ability to control their lateral COM excursion during walking had the highest scores on the BBS.

Our results also demonstrated a correlation between better lateral COM control during walking and walking speed. This relationship was moderate for preferred walking speed and strong for fast walking speed. The difference in correlation strength could be attributed to how COM excursion changes with walking speed. Studies in healthy adults reported that lateral COM excursion decreases with walking speed [53], and restrictions to COM excursion lead to increased walking speeds in order to ensure dynamic stability [54]. Thus, the ability to control lateral COM excursion could provide stability that enables individuals to walk faster.

Our findings have important clinical implications for balance assessment and training. Our study indicates that the ability to control lateral COM motion during walking is closely related to several functional measures of gait and balance in people with iSCI. We believe this relationship may be because control of lateral COM motion during walking and the gait and balance measures we examined are all dependent on the capacity of the nervous system to both anticipate and react to ongoing COM dynamics. People with iSCI have an impaired ability to anticipate COM dynamics due to sensory [55] and motor [7] dysfunction. Additionally, research has found deficits in reactive balance responses used to control lateral COM motion in this population [56]. Thus, limitations in the ability to accurately anticipate and react to ongoing COM dynamics may reduce their ability to control lateral COM motion during walking and to perform functional measures of gait and balance that require these fundamental skills. This relationship could motivate clinical interventions that directly target the ability to control lateral COM motion during walking to improve functional balance and gait in people with iSCI. The clinical interventions could focus more on traditional balance training tasks, like narrow base of support walking, rapid maneuvers during walking, or manual perturbations to improve lateral balance control. Alternately, a novel approach to train lateral COM control in people with iSCI could be to perform gait training in a movement amplification field that applies proportional forces in the same direction as the real-time lateral velocity of the participant [57, 58].

There are a few limitations associated with this study. While our findings suggest that lateral COM motion during walking is associated with clinical gait and balance outcomes, it is not clear if this relationship is causative. Future research is recommended to further investigate existence of a causal relationship between these measures. Our current study has a small sample size and relatively high functioning (community dwelling), ambulatory people with iSCI were included, thus our findings cannot be generalized to all individuals with iSCI.

## Conclusion

This study finds that the ability to control lateral COM motion during walking predicts a wide range of clinical gait and balance measures and may be a contributing factor to gait and balance outcomes in people with iSCI. Further research should explore if interventions designed to improve control of lateral COM motion during walking translate to improvements in functional gait and balance in people with iSCI.

## Supporting information

Strobe checklist

## Data Availability

The raw data supporting the conclusions of this article will be made available by the authors upon request.

## Ethics Approval

This protocol involving human participants was approved by the Institutional Review Boards at Northwestern University and the Edward Hines Jr. Veterans Affairs Hospital. The participants provided their written informed consent to participate in this study.

## Author Contributions

KG, WO, AS, TC and SD designed the study. KG, AS, HH and SD collected the data. AS, SD, KK and KG analyzed the data. All authors contributed to the manuscript and approved the submitted version.

## Acknowledgement

The authors would like to thank Shay Pinhey, Jordan Dembsky, Mackenzie Mattone, Yuchan Choi and Keri Han for their assistance in data collection as well as all the participants who took part in the study.

